# Bidirectional relationship between Sleep Disturbance and Atrial Fibrillation: a SPRINT-based analysis

**DOI:** 10.64898/2026.09.10.26362799

**Authors:** Moustafa Elnewishy, Asem M. Mohsen, Tarek Zaho, Richard Kazibwe, Takeki Suzuki, M Benjamen Shoemaker, Prashant D. Bhave, Elsayed Z. Soliman

## Abstract

**Background:** Sleep disturbance has been associated with atrial fibrillation (AF), but prior studies have largely relied on sleep assessed at a single time point or examined specific sleep disorders. Whether changes in sleep disturbance over time predict AF and, conversely, whether AF predicts subsequent sleep disturbance within the same population is not well established.

**Methods:** We conducted a secondary analysis of the Systolic Blood Pressure Intervention Trial (SPRINT), which enrolled adults with hypertension and increased cardiovascular risk without diabetes. Two prospective analytic cohorts were constructed to examine each direction of the association: 7,232 participants without baseline AF for the analysis of sleep disturbance and incident AF, and 6,079 participants without clinically significant sleep disturbance at baseline for the reciprocal analysis. Sleep disturbance was assessed repeatedly using a 0–3 patient-reported symptom score, with clinically significant sleep disturbance defined as a score ≥2. AF was ascertained from protocol 12-lead ECGs. Time-dependent Cox models evaluated time-updated sleep disturbance in relation to incident AF and time-updated AF in relation to incident clinically significant sleep disturbance, with adjustment for demographic, lifestyle, and cardiovascular risk factors.

**Results:** During a median follow-up of 3.3 years, 189 incident AF events occurred. Time-updated clinically significant sleep disturbance was associated with a 78% higher adjusted risk of incident AF (HR, 1.78; 95% CI, 1.27–2.51). Each 1-point increase in the time-updated sleep score was associated with a 26% higher risk of AF (HR, 1.26; 95% CI, 1.09–1.46). Compared with a sleep score of 0, the adjusted HRs were 1.20 (95% CI, 0.85–1.70), 2.05 (95% CI, 1.33–3.15), and 1.72 (95% CI, 1.03–2.88) for scores of 1, 2, and 3, respectively. In the reciprocal analysis, 1,422 incident clinically significant sleep-disturbance events occurred during a median follow-up of 3.2 years. Time-updated AF was associated with a 59% higher adjusted risk of incident clinically significant sleep disturbance (HR, 1.59; 95% CI, 1.16–2.16). Associations in both directions were consistent across prespecified subgroups, with no significant effect modification.

**Conclusion:** In adults with hypertension, sleep disturbance and AF were prospectively and reciprocally associated when evaluated longitudinally. These findings support a bidirectional relationship between sleep disturbance and AF and highlight the potential clinical relevance of assessing sleep health in patients with or at risk for AF.

## Introduction

Atrial fibrillation (AF) is the most common sustained cardiac arrhythmia and a major cause of stroke, heart failure, and premature mortality.^1,2^ Sleep disturbance is also highly prevalent, affecting an estimated one-third of adults in the general population, particularly those with cardiovascular risk factors, with sleep-related symptoms among the most common chronic complaints in older adults.^3,4^ Sleep disturbance and AF are especially relevant among adults with hypertension and may be linked through shared cardiovascular, autonomic, behavioral, and psychosocial pathways.^5,6^

Although sleep abnormalities have been associated with AF, most prior studies have focused on specific sleep disorders or sleep characteristics assessed at a single point in time.^5,7,8^ This approach may not adequately capture patient-perceived sleep disturbance, which is dynamic and may improve or worsen over time. Moreover, considerably less is known about the reciprocal relationship, whether AF is associated with the subsequent development of sleep disturbance. Thus, whether sleep disturbance and AF are longitudinally and bidirectionally associated when changes in both conditions are considered over time remains incompletely understood. Establishing such a relationship may have important clinical and public health implications because sleep symptoms are readily identified through patient report and represent a potentially modifiable component of cardiovascular health.

Accordingly, we examined the bidirectional longitudinal association between sleep disturbance and AF using data from the Systolic Blood Pressure Intervention Trial (SPRINT). We hypothesized that sleep disturbance would be associated with subsequent AF and that AF would, reciprocally, be associated with subsequent sleep disturbance.

## Methods

### Study Design and Population

This study was a secondary analysis of participants enrolled in the Systolic Blood Pressure Intervention Trial (SPRINT), a multicenter randomized clinical trial comparing intensive versus standard systolic blood pressure treatment. SPRINT enrolled adults aged ≥50 years with hypertension and increased cardiovascular risk but without diabetes mellitus or a history of stroke.^9,10^ The institutional review boards at all participating centers approved the original SPRINT protocol, and all participants provided written informed consent.

Of the 9,361 randomized SPRINT participants, 7,979 met the eligibility criteria for the present analysis after application of diabetes- and glucose-related exclusions, including exclusion of participants with fasting baseline glucose ≥126 mg/dL or missing fasting baseline glucose measurements. From this cohort, two separate analytic samples were constructed to evaluate each direction of the hypothesized bidirectional association. For the analysis of sleep disturbance and incident AF, participants with prevalent AF at baseline, missing baseline sleep assessment, or no follow-up ECG assessment were excluded, resulting in an eligible cohort of 7,232 participants. For the reciprocal analysis of AF and incident clinically significant sleep disturbance, participants with clinically significant sleep disturbance at baseline, missing baseline sleep assessment, or insufficient follow-up sleep or ECG data were excluded, resulting in an eligible cohort of 6,079 participants. Participants with missing data for ≥1 covariate required for the fully adjusted model were further excluded from the corresponding complete-case analyses, yielding 7,118 and 5,989 participants, respectively. (**Figure 1**).

**Figure 1.**
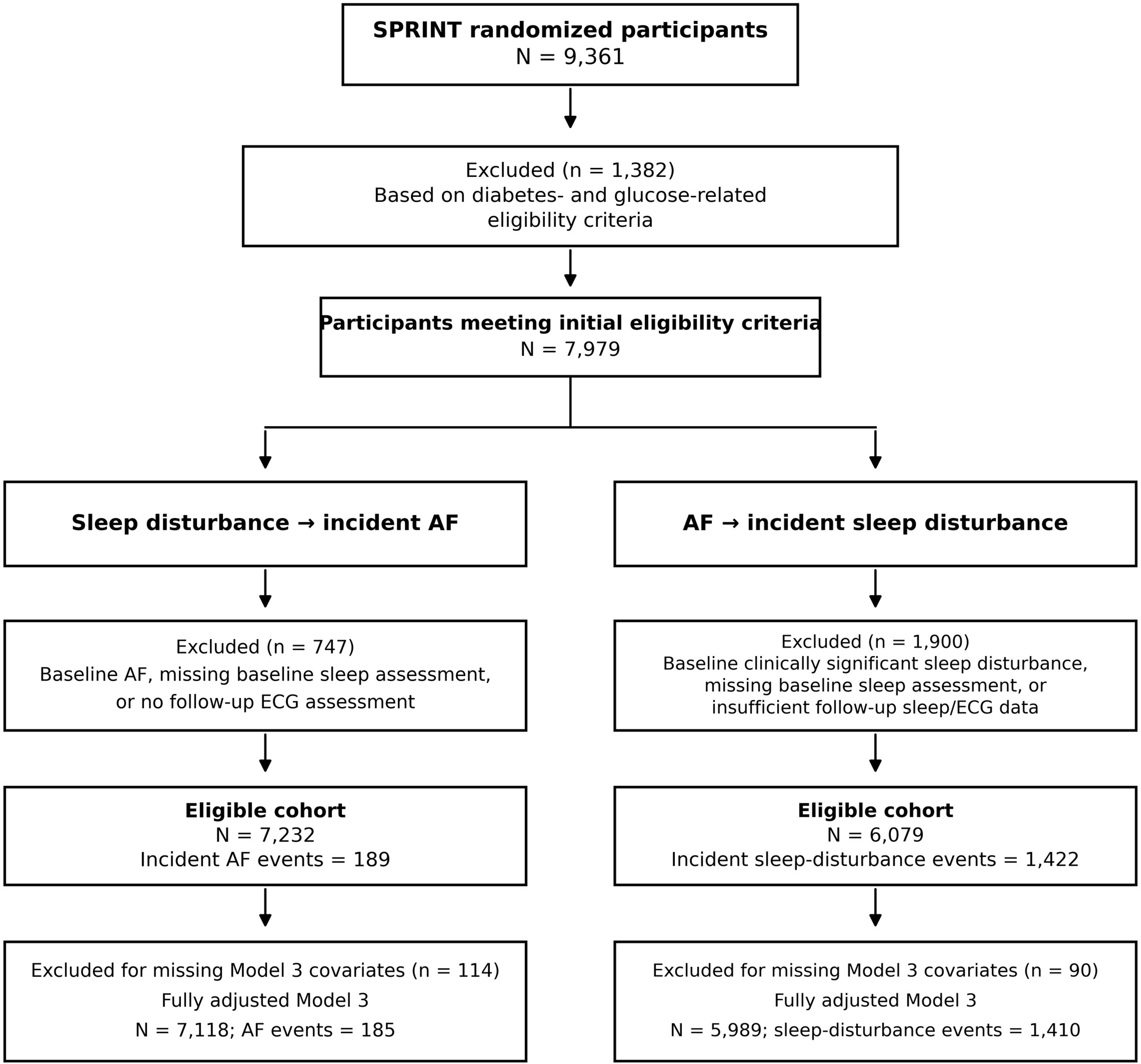
Flow of Participants Through the Study. Participant flow for the two analytic cohorts evaluating the bidirectional association between sleep disturbance and atrial fibrillation (AF). For the analysis of sleep disturbance and incident AF, participants with baseline AF, missing baseline sleep assessment, or no follow-up ECG assessment were excluded. For the reciprocal analysis of AF and incident clinically significant sleep disturbance, participants with clinically significant sleep disturbance at baseline, missing baseline sleep assessment, or insufficient follow-up sleep/ECG data were excluded. Additional participants with missing covariates were excluded from the fully adjusted Model 3 analyses. Clinically significant sleep disturbance was defined as a sleep score ≥2. AF indicates atrial fibrillation; ECG, electrocardiogram.

#### Assessment of sleep disturbance

Sleep disturbance was assessed repeatedly during follow-up using a single patient-reported item from the Patient Health Questionnaire that asked participants how often during the preceding 2 weeks they had been bothered by trouble falling or staying asleep or sleeping too much. Responses were scored from 0 (not at all) to 3 (nearly every day). Clinically significant sleep disturbance was defined a priori as a score ≥2, corresponding to symptoms occurring more than half the day or nearly every day.

#### Ascertainment of incident AF

AF was ascertained from protocol 12-lead ECGs obtained at baseline, year 2, year 4, and the closeout visit. ECGs were digitally recorded using GE MAC 1200 electrocardiographs (GE Healthcare) at standard calibration (10 mm/mV and 25 mm/s) and transmitted to the Epidemiological Cardiology Research Center (EPICARE) at Wake Forest University School of Medicine in Winston-Salem, NC.

ECG quality was reviewed by trained technicians blinded to randomized treatment assignment before automated processing using the GE Marquette 12-SL algorithm (version 2001). ECG abnormalities were classified according to the Minnesota Code.¹¹ ECGs classified as AF were visually confirmed by trained ECG readers according to previously described SPRINT procedures. AF documented on the baseline ECG was classified as prevalent AF, whereas incident AF was defined by the first follow-up protocol ECG demonstrating AF.

#### Covariates

Covariates were selected a priori based on established associations with AF and sleep disturbance and their potential to confound the associations of interest. These included age, sex, race, randomized blood pressure treatment assignment, educational attainment, smoking status, alcohol consumption, physical activity, body mass index, systolic blood pressure, number of antihypertensive medications, prevalent cardiovascular disease, total cholesterol, high-density lipoprotein cholesterol, triglycerides, estimated glomerular filtration rate, statin use, and aspirin use. These variables were assessed at baseline.

### Statistical Analysis

Baseline characteristics were summarized according to the occurrence of incident AF in the sleep-to-AF cohort and incident clinically significant sleep disturbance in the AF-to-sleep cohort. Continuous variables are presented as mean ± SD or median (interquartile range), as appropriate, and categorical variables as number (percentage). Between-group differences were evaluated using analysis of variance or the Kruskal-Wallis test for continuous variables and the χ² test for categorical variables, as appropriate.

#### Sleep Disturbance and Incident AF

The association between sleep disturbance and incident AF was evaluated using Cox proportional hazards regression with sleep disturbance modeled as a time-updated exposure. Follow-up for each participant was partitioned into intervals defined by successive sleep assessments. At the beginning of each interval, the most recently available sleep score was assigned and carried forward until the next assessment, incident AF, censoring, or end of follow-up. Participants could therefore move between sleep categories over time according to their current reported sleep status.

Sleep disturbance was examined using three complementary time-updated parameterizations. First, it was modeled as a binary exposure, with scores ≥2 representing clinically significant sleep disturbance and scores <2 serving as the reference. Because sleep status was updated at each assessment, participants could contribute person-time to either category and could move in either direction as their reported symptoms changed. Second, the 0-to-3 sleep score was modeled as a continuous time-updated variable, with the HR representing the relative difference in the hazard of incident AF associated with each 1-point higher current sleep score. Third, sleep score was modeled as a four-level time-updated categorical variable (0, 1, 2, and 3), with score 0 as the reference. At each follow-up interval, participants contributed person-time according to their current sleep score. Accordingly, the HRs for scores 1 versus 0, 2 versus 0, and 3 versus 0 compared the hazard of incident AF during person-time characterized by the respective current sleep score with person-time characterized by a current score of 0. Participants were allowed to transition among all four categories during follow-up. A global Wald test assessed the overall association across sleep-score categories.

As a sensitivity analysis, the same binary, continuous, and categorical parameterizations were evaluated using baseline sleep status only. In these models, baseline sleep exposure was treated as fixed throughout follow-up, regardless of subsequent changes in sleep symptoms.

#### AF and Incident Sleep Disturbance

For the reciprocal analysis, participants with clinically significant sleep disturbance at baseline were excluded, and the association between AF and incident clinically significant sleep disturbance was evaluated using time-dependent Cox proportional hazards regression. AF was modeled as a time-updated exposure incorporating both prevalent and incident AF. Participants with AF at baseline contributed all follow-up time as AF-exposed. Participants without baseline AF contributed person-time to the non-AF group until AF was first documented and, after AF detection, contributed all subsequent person-time to the AF-exposed group. Unlike sleep disturbance, AF status was therefore treated as an absorbing exposure state; once AF was identified, participants did not revert to the non-AF category.

The outcome was the first occurrence of clinically significant sleep disturbance (sleep score ≥2). The resulting HR compared the instantaneous hazard of developing clinically significant sleep disturbance during AF-exposed versus AF-unexposed person-time.

As a sensitivity analysis, AF was restricted to baseline AF and treated as a fixed exposure throughout follow-up, irrespective of subsequent incident AF.

#### Multivariable Models

For both directions of association, three sequential Cox models were fitted. Model 1 was unadjusted. Model 2 adjusted for age, sex, race, and randomized treatment assignment. Model 3 additionally adjusted for education, smoking status, physical activity, alcohol use, body mass index, systolic blood pressure, number of antihypertensive medications, prevalent cardiovascular disease, total cholesterol, HDL cholesterol, triglycerides, estimated glomerular filtration rate, statin use, and aspirin use. HRs and 95% CIs were reported. The eligible cohorts were used for the unadjusted analyses, whereas fully adjusted analyses were restricted to participants with complete covariate data.

#### Cumulative Incidence and Subgroup Analyses

Because the primary exposures varied over time, cumulative incidence was illustrated using Simon–Makuch curves, which accommodate transitions in time-dependent exposure status. Differences between curves were evaluated using the Mantel–Byar test, the time-dependent analogue of the log-rank test, and numbers at risk were displayed below the curves.

Prespecified subgroup analyses evaluated the consistency of the primary time-updated associations according to age (<75 versus ≥75 years), sex, race (Black versus non-Black), prevalent cardiovascular disease, and randomized blood pressure treatment assignment. Fully adjusted Cox models were fitted within each subgroup, and effect modification was evaluated by multiplicative interaction terms using Wald tests. Subgroup-specific HRs and 95% CIs, together with the numbers of participants and events, were displayed in forest plots.

All statistical tests were 2-sided, with P<0.05 considered statistically significant. Analyses were performed using SAS version 9.4 (SAS Institute Inc).

## Results

### Study Population and Baseline Characteristics

Participant selection for the two analytic cohorts is shown in Figure 1. The sleep disturbance-to-AF analysis included 7,232 participants, among whom 189 incident AF events occurred. After exclusion of 114 participants with missing covariate data, 7,118 participants with 185 incident AF events were included in the fully adjusted analysis. Median follow-up was 3.3 years. The reciprocal AF-to-sleep disturbance analysis included 6,079 participants, among whom 1,422 developed incident clinically significant sleep disturbance. After exclusion of 90 participants with missing covariate data, 5,989 participants with 1,410 events were included in the fully adjusted analysis. Median follow-up was 3.2 years.

Baseline characteristics according to incident AF and incident clinically significant sleep disturbance are shown in **Table 1**. Participants who developed AF were older, less frequently Black, had lower diastolic blood pressure and estimated glomerular filtration rate, used more antihypertensive medications, and more frequently had prevalent cardiovascular disease than those who did not develop AF. Baseline sleep score and the prevalence of clinically significant sleep disturbance were similar between participants who did and did not subsequently develop AF. Participants who developed clinically significant sleep disturbance were more frequently women and Black, had fewer years of education, were more frequently current smokers, reported less vigorous physical activity, had higher body mass index, and more frequently had prevalent cardiovascular disease than those who did not develop clinically significant sleep disturbance

**Table 1.**
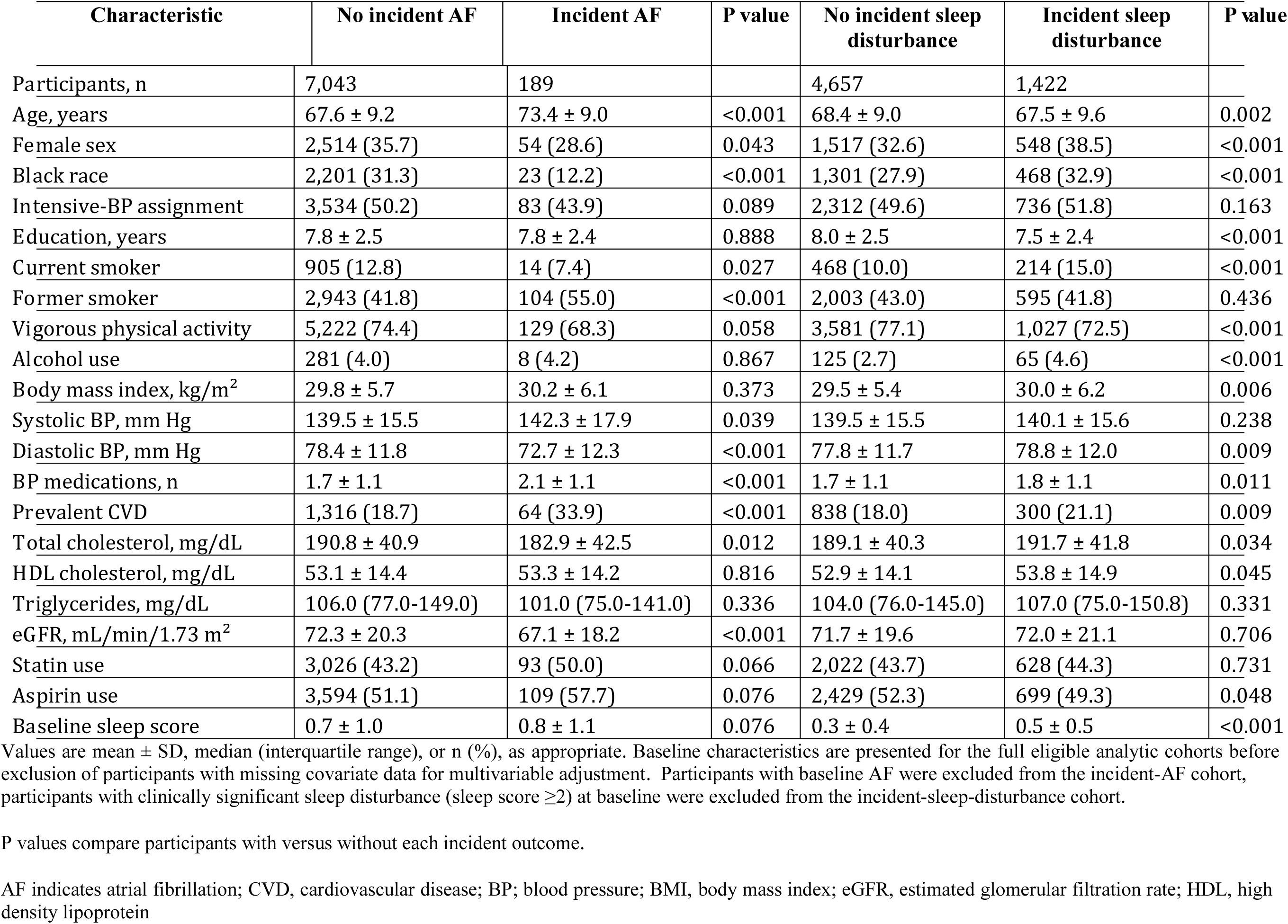
Baseline Characteristics According to Incident AF and Incident Sleep Disturbance.

### Sleep Disturbance and Incident AF

Time-updated clinically significant sleep disturbance (sleep score ≥2 versus <2) was associated with a higher risk of incident AF (**Table 2**). The association was present in the unadjusted model (HR, 1.74; 95% CI, 1.25–2.42; P=0.001) and remained significant after full multivariable adjustment (HR, 1.78; 95% CI, 1.27–2.51; P<0.001).

**Table 2.**
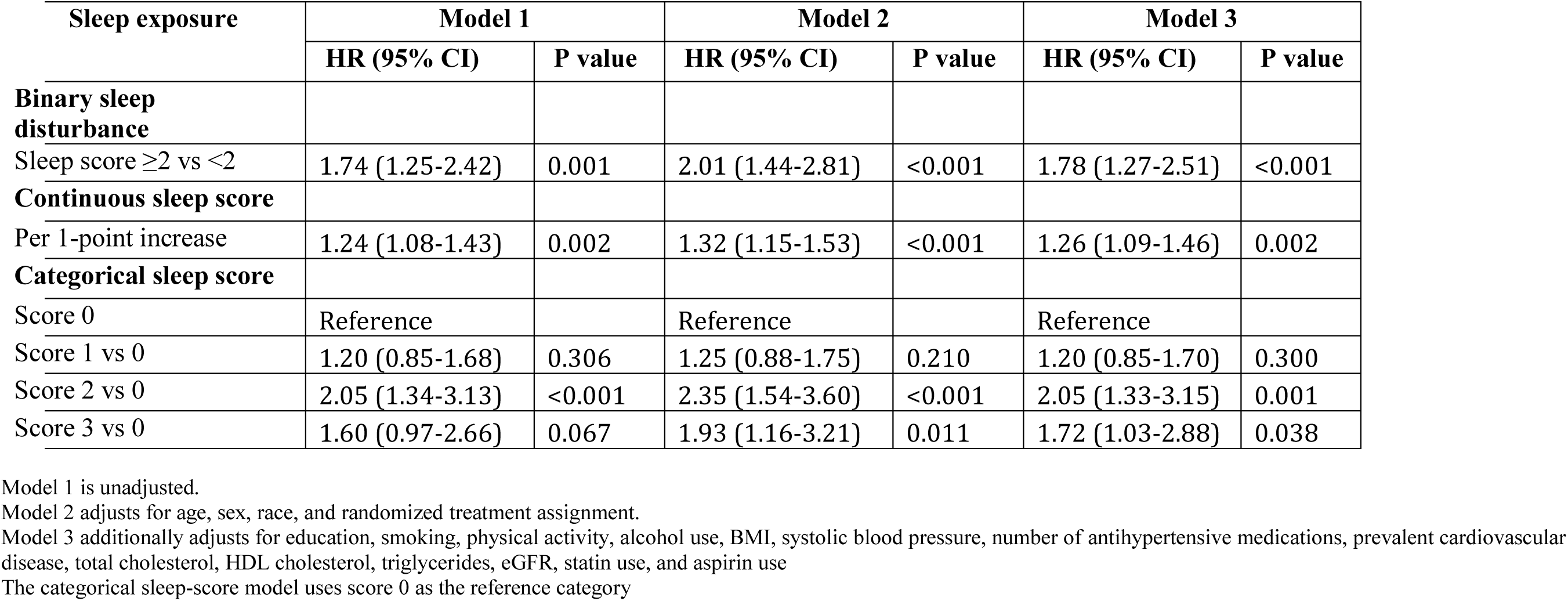
Association Between Time-Updated Sleep Disturbance and Incident Atrial Fibrillation.

The association was also evident when the full range of sleep scores was considered. Each 1-point increase in the time-updated sleep score was associated with a 26% higher risk of incident AF after full adjustment (HR, 1.26; 95% CI, 1.09–1.46; P=0.002). When modeled categorically, compared with a current sleep score of 0, the fully adjusted HRs for incident AF were 1.20 (95% CI, 0.85–1.70; P=0.300) for a score of 1, 2.05 (95% CI, 1.33–3.15; P=0.001) for a score of 2, and 1.72 (95% CI, 1.03–2.88; P=0.038) for a score of 3. The overall association across sleep-score categories was significant (global P=0.006).

In sensitivity analyses using baseline rather than time-updated sleep measures, associations were generally attenuated (**Supplementary Table 1**). For clinically significant sleep disturbance, the fully adjusted association was directionally consistent but was not statistically significant (HR, 1.33; 95% CI, 0.93–1.89; P=0.114). Baseline sleep score remained associated with incident AF when modeled continuously (HR per 1-point increase, 1.17; 95% CI, 1.01–1.34; P=0.032), and a baseline score of 3 was associated with higher AF risk compared with a score of 0 (HR, 1.71; 95% CI, 1.09–2.68; P=0.020). However, the overall association across baseline sleep-score categories was not statistically significant (global P=0.135).

Consistent with the Cox regression findings, Simon–Makuch curves demonstrated a higher cumulative incidence of AF during follow-up among participants with time-updated clinically significant sleep disturbance than among those without clinically significant sleep disturbance (Mantel–Byar P<0.001; **Figure 2A**).

**Figure 2.**
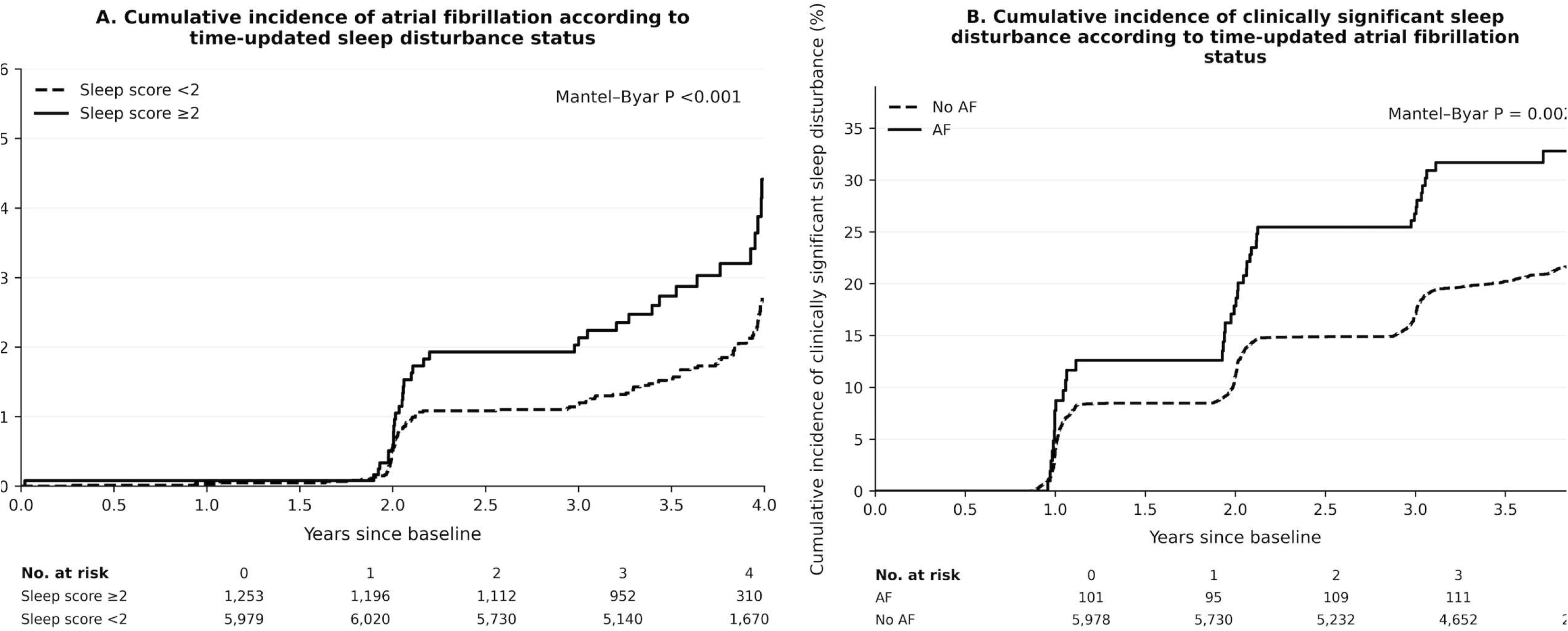
Cumulative Incidence of Atrial Fibrillation and Sleep Disturbance According to Time-Updated Exposure Status. Simon–Makuch curves showing cumulative incidence according to time-updated exposure status. **A**, Cumulative incidence of atrial fibrillation according to time-updated clinically significant sleep disturbance status (sleep score ≥2 vs <2). **B**, Cumulative incidence of clinically significant sleep disturbance according to time-updated atrial fibrillation status. Because exposure status could change during follow-up, Simon–Makuch curves were used to account for transitions between exposure groups. Differences between curves were evaluated using the Mantel–Byar test. Numbers at risk are shown below each panel. AF indicates atrial fibrillation.

### AF and Incident Sleep Disturbance

In the reciprocal analysis, time-updated AF was associated with a higher risk of incident clinically significant sleep disturbance (Table 3). The unadjusted HR was 1.59 (95% CI, 1.18–2.15; P=0.002). After full multivariable adjustment, AF remained associated with a 59% higher risk of incident clinically significant sleep disturbance (HR, 1.59; 95% CI, 1.16–2.16; P=0.004).

**Table 3.** Association Between Atrial Fibrillation and Incident Clinically Significant Sleep Disturbance.

| AF exposure | Model 1 |  | Model 2 |  | Model 3 |  |
| --- | --- | --- | --- | --- | --- | --- |
|  | HR (95% CI) | P value | HR (95% CI) | P value | HR (95% CI) | P value |
| Time-updated AF (baseline + incident) | 1.59 (1.18-2.15) | 0.002 | 1.76 (1.30-2.38) | <0.001 | 1.59 (1.16-2.16) | 0.004 |
| Baseline AF only | 1.43 (0.99-2.05) | 0.054 | 1.58 (1.09-2.27) | 0.015 | 1.45 (1.00-2.10) | 0.047 |

Model 1 is unadjusted.
Model 2 adjusts for age, sex, race, and randomized treatment assignment.
Model 3 additionally adjusts for education, smoking, physical activity, alcohol use, BMI, systolic blood pressure, number of antihypertensive medications, prevalent cardiovascular disease, total cholesterol, HDL cholesterol, triglycerides, eGFR, statin use, and aspirin use

When AF was restricted to baseline AF and treated as a fixed exposure, the association remained significant after full adjustment (HR, 1.45; 95% CI, 1.00–2.10; P=0.047).

Simon–Makuch curves similarly demonstrated a higher cumulative incidence of clinically significant sleep disturbance during AF-exposed than AF-unexposed follow-up (Mantel–Byar P=0.002; **Figure 2B**).

### Subgroup Analyses

The associations in both directions were generally consistent across prespecified subgroups (Figure 3). The association between time-updated clinically significant sleep disturbance and incident AF did not significantly differ by age, sex, race, prevalent cardiovascular disease, or randomized blood pressure treatment assignment (all P for interaction >0.05; **Figure 3A**). Similarly, there was no significant effect modification of the association between time-updated AF and incident clinically significant sleep disturbance across any of these subgroups (all P for interaction >0.05; **Figure 3B**).

**Figure 3.**
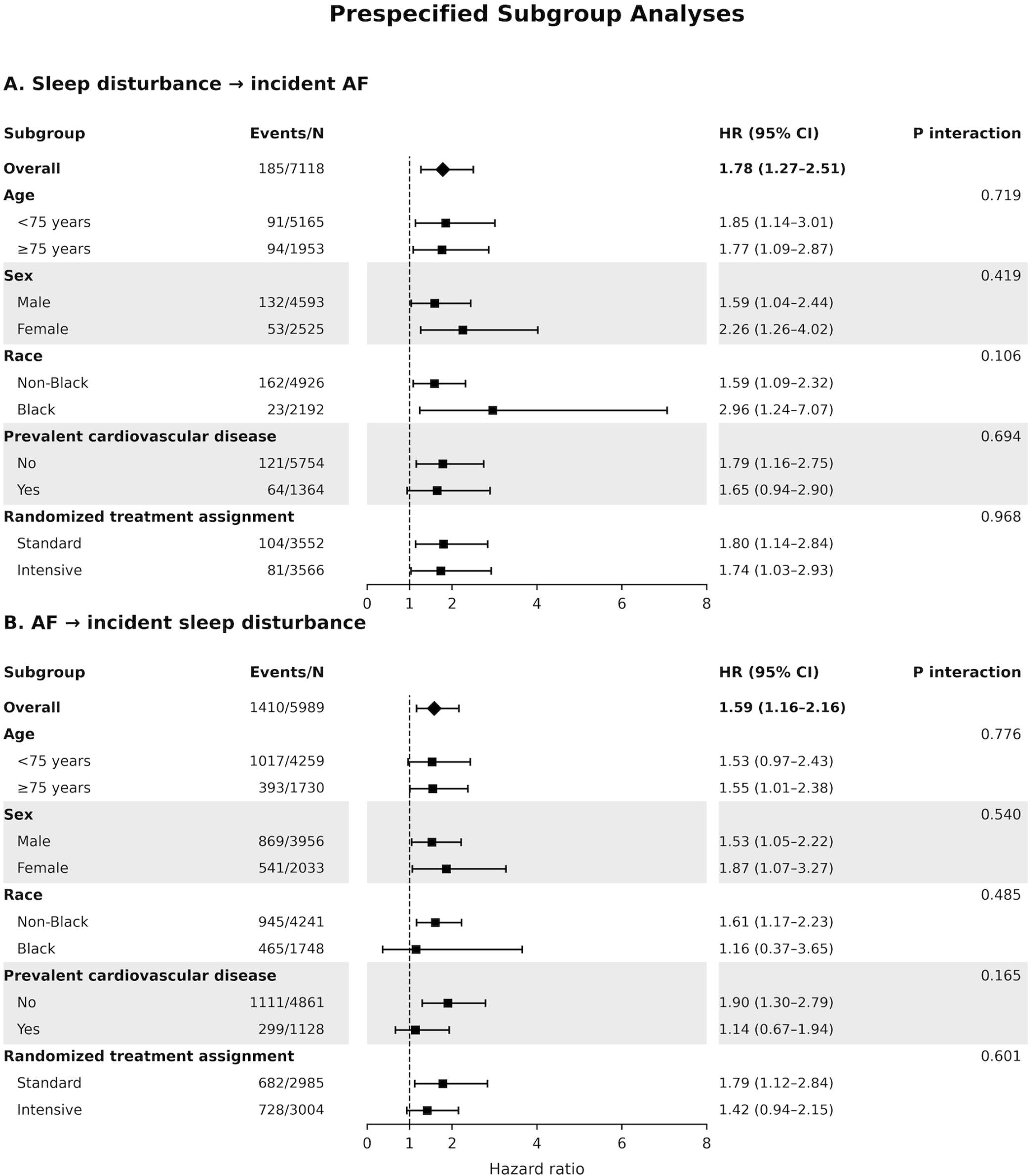
Subgroup Analyses of the Bidirectional Associations Between Sleep Disturbance and Atrial Fibrillation. Forest plots showing fully adjusted hazard ratios (HRs) and 95% confidence intervals (CIs) for the primary time-updated associations across prespecified subgroups. **A**, Association of time-updated clinically significant sleep disturbance with incident atrial fibrillation. **B**, Association of time-updated atrial fibrillation with incident clinically significant sleep disturbance. Subgroups were defined by age (<75 vs ≥75 years), sex, race, prevalent cardiovascular disease, and randomized blood pressure treatment assignment. Models were adjusted for age, sex, race, randomized treatment assignment, educational attainment, smoking status, physical activity, alcohol consumption, body mass index, systolic blood pressure, number of antihypertensive medications, prevalent cardiovascular disease, total cholesterol, high-density lipoprotein cholesterol, triglycerides, estimated glomerular filtration rate, statin use, and aspirin use, with the subgroup-defining variable omitted from the corresponding stratified model. *P* values for interaction were obtained from multiplicative interaction terms. AF indicates atrial fibrillation; CI, confidence interval; HR, hazard ratio.

## Discussion

In this secondary analysis of SPRINT participants with hypertension and without diabetes, we found evidence of a bidirectional longitudinal association between patient-reported sleep disturbance and AF. Time-updated clinically significant sleep disturbance was associated with a 78% higher risk of incident AF after adjustment for demographic, lifestyle, and cardiovascular risk factors. The association was also evident when sleep disturbance was evaluated continuously and categorically: each 1-point increase in the time-updated sleep score was associated with a 26% higher risk of AF, and sleep scores of 2 and 3 were each associated with higher AF risk compared with a score of 0. In the reciprocal direction, time-updated AF was associated with a 59% higher risk of subsequent clinically significant sleep disturbance. These associations were generally consistent across prespecified subgroups. Collectively, these findings suggest that the relationship between sleep disturbance and AF may extend in both directions and underscore the importance of considering sleep symptoms as a dynamic rather than exclusively baseline characteristic.

Our finding that sleep disturbance is associated with incident AF adds to a growing literature linking abnormal sleep with AF.⁵ However, previous studies have largely evaluated specific sleep disorders or sleep characteristics assessed at a single point in time.^5,7,8^ The present findings extend this literature by incorporating repeated patient-reported sleep assessments and allowing sleep status to vary during follow-up. This distinction may be important because sleep symptoms fluctuate over time, and a single baseline measurement may incompletely characterize an individual’s sleep experience. Consistent with this possibility, the association of clinically significant sleep disturbance with AF was attenuated when sleep was restricted to baseline status (HR, 1.33; 95% CI, 0.93–1.89) compared with the time-updated analysis (HR, 1.78; 95% CI, 1.27–2.51). Similarly, the association per 1-point increase in sleep score was numerically weaker in the baseline-only analysis (HR, 1.17; 95% CI, 1.01–1.34) than in the time-updated analysis (HR, 1.26; 95% CI, 1.09–1.46). Although these estimates were not formally compared, the findings suggest that incorporating changes in sleep symptoms over time may provide information not captured by a single baseline assessment.

The complementary parameterizations of sleep disturbance provide additional insight. Each 1-point increase in the current sleep score was associated with a 26% higher risk of AF, while categorical analyses demonstrated significantly higher risks among participants with sleep scores of 2 and 3 compared with those reporting no symptoms. The association was strongest for a score of 2 (HR, 2.05; 95% CI, 1.33–3.15), while a score of 3 was associated with a 72% higher risk (HR, 1.72; 95% CI, 1.03–2.88). Thus, although the findings support an association across increasing frequencies of sleep symptoms, the categorical point estimates did not demonstrate a strictly monotonic dose-response pattern. Importantly, the sleep measure captured patient-reported difficulty initiating or maintaining sleep or sleeping too much, rather than a specific sleep disorder. Patient-reported symptoms may be clinically informative because they reflect the sleep problems experienced by patients and are readily elicited in routine clinical care. At the same time, these findings should not be interpreted as establishing associations with specific conditions such as insomnia or obstructive sleep apnea (OSA).

Several mechanisms could plausibly link disturbed sleep with AF. Sleep disruption has been associated with autonomic imbalance, including increased sympathetic activity and altered parasympathetic regulation, which may influence atrial electrophysiology and susceptibility to arrhythmia.^12^ Sleep disturbance has also been associated with systemic inflammation, oxidative stress, and neurohormonal activation, pathways implicated in atrial structural and electrical remodeling.¹² Related mechanisms are particularly well characterized in OSA, in which intermittent hypoxemia, intrathoracic pressure changes, and recurrent autonomic surges may promote an arrhythmogenic atrial substrate.^13^ Although OSA was not assessed in the present analysis and may partly contribute to the observed association, the consistency of the association when sleep disturbance was modeled using binary, continuous, and categorical parameterizations supports the robustness of the overall finding.

An important and less well studied finding was the reciprocal association of AF with subsequent sleep disturbance. Time-updated AF was associated with a 59% higher adjusted risk of developing clinically significant sleep disturbance. AF may adversely affect sleep through several pathways, including nocturnal palpitations, heightened awareness of an irregular heartbeat, dyspnea, and the psychological burden associated with an unpredictable recurrent arrhythmia.^6^ Anxiety regarding AF episodes and recurrence may further interfere with sleep initiation and maintenance, potentially creating a cycle in which sleep disturbance and AF reinforce one another.^6^ The association was also observed when AF was restricted to baseline status (HR, 1.45; 95% CI, 1.00–2.10), supporting the overall finding using an alternative exposure definition. The somewhat larger point estimate in the time-updated analysis should be interpreted cautiously because the baseline and time-updated estimates were not formally compared.

These findings may have clinical implications. Sleep health is included in the American Heart Association’s Life’s Essential 8 construct, reflecting increasing recognition of sleep as a component of cardiovascular health.^14^ Our findings extend this concept to AF by suggesting that recurrent or evolving sleep symptoms may identify hypertensive adults at increased risk for AF, while AF may in turn identify patients at increased risk of developing clinically meaningful sleep complaints. Because patient-reported sleep symptoms can be assessed readily in routine practice, incorporating questions about sleep into the clinical evaluation of patients at risk for or living with AF may provide useful information. However, our findings do not establish that screening for AF solely on the basis of sleep disturbance improves outcomes, nor do they establish that treating sleep disturbance prevents AF. Whether interventions directed at sleep—including behavioral approaches or evaluation and treatment of specific sleep disorders when clinically indicated—can reduce AF incidence or burden warrants prospective investigation.

This study has several strengths. SPRINT provided a large, well-characterized cohort of adults with hypertension, standardized phenotyping, and protocol-based ECG ascertainment with centralized interpretation. The bidirectional design used separate analytic cohorts in which participants with the outcome of interest at baseline were excluded, allowing the temporal association to be evaluated in both directions. Repeated assessment of patient-reported sleep symptoms permitted sleep to be modeled dynamically rather than solely at baseline, while time-updated AF status incorporated AF developing during follow-up. Evaluation of sleep disturbance using binary, continuous, and categorical parameterizations demonstrated generally consistent associations across alternative representations of the exposure, and the findings were generally consistent across clinically relevant subgroups.

Several limitations should also be considered. First, this was a secondary observational analysis of a randomized clinical trial, and causality cannot be inferred despite the longitudinal design and multivariable adjustment. Second, sleep disturbance was assessed using a single patient-reported symptom item rather than a dedicated sleep questionnaire, actigraphy, polysomnography, or clinical sleep evaluation. The measure therefore captures perceived sleep disturbance but cannot distinguish among specific sleep disorders or characterize sleep architecture, duration, or sleep-disordered breathing. This limitation is balanced in part by the availability of repeated assessments, which allowed changes in symptom burden to be incorporated over time. Third, OSA was not systematically assessed and could represent an important source of residual confounding. Other unmeasured or incompletely measured factors, including psychological symptoms, medication effects, and changes in health status, may also contribute to both sleep disturbance and AF. Fourth, AF ascertainment based on intermittent protocol ECGs may have missed paroxysmal or asymptomatic AF, potentially resulting in exposure or outcome misclassification. Finally, SPRINT enrolled adults aged ≥50 years with hypertension and increased cardiovascular risk while excluding individuals with diabetes or prior stroke; therefore, the findings may not generalize to other populations such as younger, lower-risk populations or to patients excluded from SPRINT.

### Conclusion

Among adults with hypertension and without diabetes, time-updated patient-reported sleep disturbance was associated with subsequent AF, and time-updated AF was reciprocally associated with subsequent clinically significant sleep disturbance. The association between sleep disturbance and AF was consistently observed when sleep symptoms were evaluated using binary, continuous, and categorical measures and was generally consistent across clinically relevant subgroups. These findings support a longitudinal, bidirectional relationship between sleep disturbance and AF and highlight the potential clinical relevance of assessing sleep symptoms in patients at risk for or living with AF. Further studies are needed to determine whether interventions targeting sleep disturbance can modify AF risk and whether AF-directed treatment can improve subsequent sleep health.

## Acknowledgments

Data for this study were obtained from the publicly available SPRINT data repository through the National Heart, Lung, and Blood Institute (NHLBI) Biologic Specimen and Data Repository Information Coordinating Center (BioLINCC). The present analysis was conducted independently of the SPRINT Research Group, and the conclusions reported herein do not necessarily represent the views of the SPRINT investigators or the NHLBI.

## Availability of data and materials

The SPRINT data used in this analysis are publicly available through the National Heart, Lung, and Blood Institute (NHLBI) Biologic Specimen and Data Repository Information Coordinating Center (BioLINCC) at https://biolincc.nhlbi.nih https://biolincc.nhlbi.nih.gov/studies/sprint/

---

**Supplementary Table 1.**
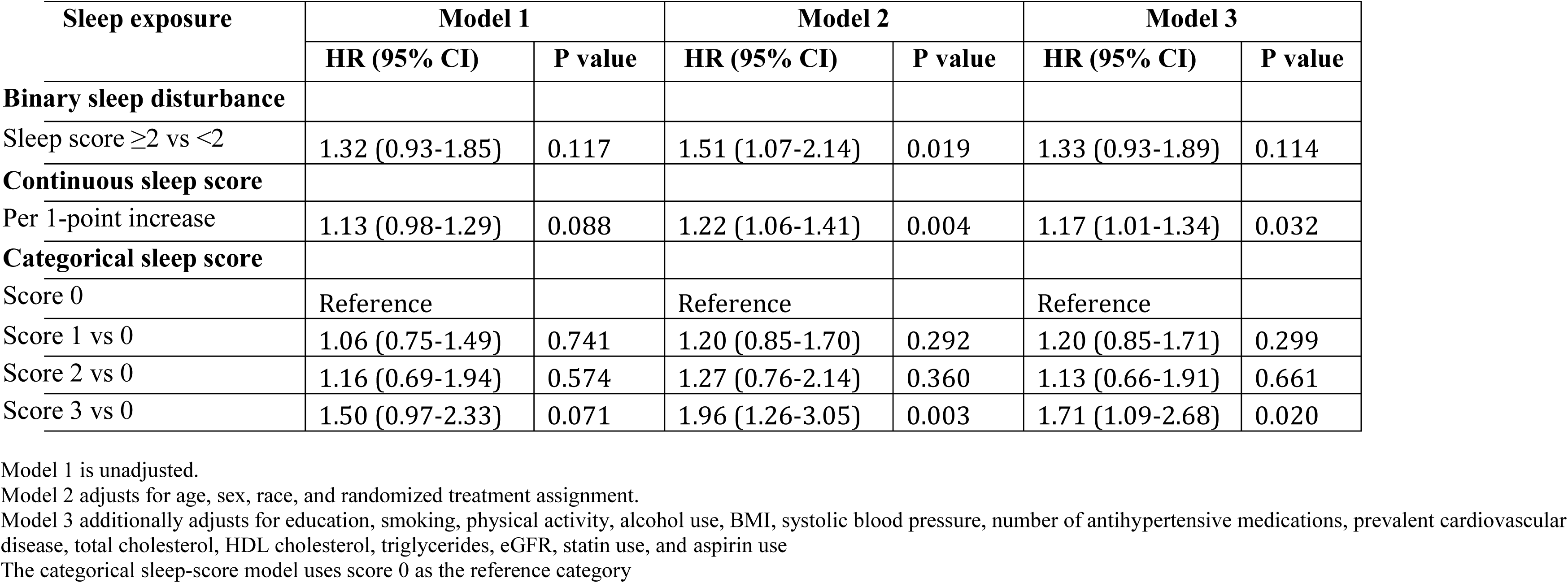
Association Between Baseline Sleep Disturbance and Incident Atrial Fibrillation.

